# Bacteriological Quality of Beef and Hygiene Practices of Food Handlers in Butcheries in Kasama District, Zambia

**DOI:** 10.1101/2020.06.06.20124214

**Authors:** Sara Hanyinza, Kunda Ndashe, Ruth Mfune, Emmanuel Chirwa, Grace Mwanza, Bruno Phiri, Bernadette Mumba, Maron Mubanga, Bernard Hang’ombe

## Abstract

The most common health risk associated with consumption of beef is microbial contamination, therefore the study was aimed to assess the level of bacterial contamination of beef and evaluate the hygienic practices in butcheries in Kasama district. Beef samples were collected from participating butcheries and a structured questionnaire was also administered to the food handlers.

Microbiological quality of the beef samples was determined by Aerobic Plate Count (APC), Faecal Coliform Count (FCC) and bacterial isolation such as *Salmonella* spp and *Staphylococcus aureus*. The APC results revealed 40.7% of the butcheries sold meat in good bacterial condition (<4 Log10 cfu/g) while 40.7% and 18.5% were critical (4-5 Log10 cfu/g) and non-acceptable conditions (>5 Log10 cfu/g), respectively. The FCC revealed that 74.1% of the butcheries sold meat in good bacterial conditions (<2 Log10 cfu/g), while 14.8% and 11.1% were critical (2-3 Log10 cfu/g) and non-acceptable conditions (>3 Log10 cfu/g), respectively. *Staphylococcus aureus* was isolated from 37% of the butcheries, none of the outlets recorded *Salmonella* spp.

Overall, the microbial quality of most (74.1%) of the market ready beef in Kasama district was acceptable for human consumption. Therefore, regular bacteriological monitoring and maintaining hygiene in the sales outlets and distribution chain is mandatory.

## 1. INTRODUCTION

Meat and meat products in many parts of the world constitutes a bigger part of a typical human diet for reasons such as health, economy and culture (Pighin et al., 2016). It is a known fact that Beef contain protein, lipids, vitamins and trace elements and is an important source of proteins to humans and it is very popular in Zambia among many households who can afford it. Beef can be accessed at retail shops, butcheries, selected farms and open markets and can be a potential source of many pathogens such as bacteria (Zhang et al., 2010). It is a very perishable commodity because of its rich nutrients that supports microbial growth (Ukut et al., 2010). The water activity of beef, approximately 0.99, is suitable for microbial growth thereby supporting proliferation of bacteria that attach and establish themselves on meat (Kinsella et al., 2006). The microbiological contamination of carcasses occurs mainly during removal of hides, evisceration, processing, packaging and storage and distribution at slaughter houses and retailed outlets (Abdalla et al., 2009).

Microorganisms that contaminate meat do not only predispose it to spoilage but are also frequently implicated in the spread of foodborne illness (Humphrey et al., 2007). During slaughter and processing, all potentially edible tissues are subjected to contamination from a variety of sources within and outside the animal (Datta et al., 2012). The pathogenic microorganisms that are implicated in contaminating meat and its products include; *Salmonella* spp., *Shigella* spp., *Campylobacter jejuni, Campylobacter coli, Yersinia enterocolitica*, verotoxigenic *Escherichia coli* (*E. coli*) and *Listeria monocytogenes* (Little et al., 2008; Meyer et al., 2010; Warren et al., 2007). *Staphylococcus aureus* contaminates beef through unhygienic handling of the meat and its products by butchery staff as this organism is a normal flora on the skin of humans (Kebede et al., 2016). Contamination of beef with *Salmonella* spp., *Campylobacter jejuni, Campylobacter coli, Yersinia enterocolitica*, and verotoxigenic *Escherichia coli* (*E. coli*) is an indication poor evisceration (Nouichi and Hamdi, 2009) while *Shigella* spp being found in gastrointestinal tract of human indicates poor hand hygiene among the handlers (Ahmed and Shimamoto, 2014). Meat and their products when contaminated can serve as vehicles of pathogens to consumers (Bhandare et al., 2007) and also reduces the shelf life of the product (Nychas et al., 2008). Several researchers have reported disease outbreaks due to consumption of contaminated beef and by products (Kadariya et al., 2014; Mead et al., 2006; Newell et al., 2010). Some diseases such as shigellosis (dysentery), salmonellosis, gastroenteritis (due to *E. coli*), and staphylococcus intoxication have been attributed to the consumption of contaminated beef (DuPont, 2007).

Monitoring levels and presence of microorganisms in meat is an important step in good management practice of butcheries and beef value chain (Poumeyrol et al., 2010). Potential safety and quality in raw meat products can be estimated with the use of indicator microorganisms including aerobic plate count (APC), coliform count (CC), *E. coli* count (ECC) (Kim and Yim, 2016). Aerobic plate count provides an estimation of the total bacterial population, therefore higher APC usually relates to poorer quality of meat and reduced shelf life (McCain et al., 2015). Coliform count provides an estimation of faecal contamination and poor sanitation during the processing of raw beef (Al-Mutairi, 2011). High CC generally correlates with higher levels of foodborne pathogens of faecal origin (Milios et al., 2014).

In Zambia, a number of studies have been conducted to assess the bacteriological quality of poultry meat (Chishimba et al., 2016; Shamaila et al., 2018; William et al., 2012). However, there are limited reports on the microbiological quality of raw beef that is market ready. This study was conducted to assess the level of bacterial contamination of beef and furthermore evaluate the hygienic practices of butcheries in Kasama District in northern Zambia.

## 2. MATERIALS AND METHODS

### 2.1. Study design and sample population

A cross-sectional study was conducted from March to April 2017 in Kasama district, Northern Province of Zambia. Kasama district is the provincial capital of the Northern Province with an estimated population of 231,824 people (2010 Census of Population National Analytical Report). The study population included 27 butcheries which were legally registered to the local municipality during the time of the research. These 27 butcheries were located in 4 townships mainly Mulilansolo, Lukupa, Kupumaula and Buseko.

In order to estimate the number of raw meat sample to collect for the study the formula below was used:

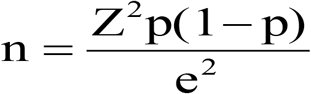

Where, n = sample size, Z = score (1.96 at 95% confidence level), P= prevalence of beef contamination in previous study [19%; (Zhao et al., 2001)], e = margin error (0.05). We calculated 161 meat samples and therefore 6 samples were collected from each of the 27 butcheries.

### 2.2. Sample and data collection

Each beef sample weighed approximately 100g and were aseptically removed and placed in a sterile sample bag and frozen immediately. The beef was collected from display refrigerator of the participating butcheries and transported to the University of Zambia, school of Veterinary Medicine, Paraclinical Sciences bacteriology laboratory within 24 hrs of collection.

Furthermore, questionnaires seeking information on the source of the beef, hygienic practices and basic meat management in the abattoir were administered to the personnel managing the participating butcheries.

### 2.3. Microbial analysis

At the laboratory, 25 gram of each meat sample was weighed out, ground and then homogenized in 225 ml of phosphate buffer saline. The homogenates were used for all the microbiological analyses.

#### 2.3.1. Determination of Total and Faecal Coliform Count

One mL of the homogenate was serially diluted in an aseptic condition and used for the enumeration of microorganisms. Ten-fold dilutions of the homogenates were made as described by Fawole and Oso, (2001). The serially diluted homogenates of the meat samples were inoculated on aerobic plate agar (Himedia, India) by pour plate method for total coliform count (TCC) and MacConkey agar (Himedia) for faecal coliform counts (FCC). The plates were incubated at 37°C aerobically for 24 hours.

The mean number of colonies counted for all count types was expressed as log10 colony forming units per gram (cfu g-1) of sample. The expressions “good condition, critical condition, unacceptable” were used to determine quality of the samples according to the microbiological criteria for fresh beef meat of the Food and Agriculture Organisation (FAO) (Heinz and Hautzinger, 2007).

#### 2.3.2. Isolation of Pathogens

##### 2.3.2.1. *Salmonella* spp

One ml of each initial homogenate of meat sample was thoroughly mixed in 9ml buffered peptone water (Himedia, India) and incubated for 20 hours at 37° C as pre-enrichment for Salmonella. From each pre-enriched sample, 1 ml of the pre-enriched sample was used to inoculate 10 ml of the selenite-cysteine medium (Himedia, India) and incubated for 7 hours at 37° C (SC). Bacterial isolation was achieved on xylose lysine deoxycholate (XLD) (Himedia, India) at 37° C for 24 hours. The colony characteristics were observed and Gram staining was performed and bacteria that were identified as presumptive Salmonella spp were further inoculated Triple sugar iron agar (TSI) agar (Oxoid, UK) and SIM agar (Oxoid, UK), which were incubated at 37° C for 18hours. Other biochemical tests for the salmonella spp isolates included; Urease, citrate and Methyl Red Voges Proskauer (MRVP) media (Oxoid, UK).

##### 2.3.2.2. Staphylococcus aureus

The meats homogenates were mixed with equal volumes (60ml) of buffered peptone water (Himedia, India) and thoroughly mixed for 5min. A50-ml aliquot of the mixture was enriched in an equal volume of double-strength enrichment broth (Trypticase soy broth supplemented with 10% NaCl and 1% sodium pyruvate). After 24 hours of incubation at 35°C, the enrichment broth was streaked on Baird-Parker (Himedia, India). Following 48hours of incubation, three to six presumptive Staphylococcus aureus (black colonies surrounded by 2-to 5-mm clear zones) were transferred to Trypticase soy agar plates followed by confirmation by a tube coagulase test (Remel, Lenexa, KS).

## 3. RESULTS

### 3.1. Bacteriological quality of beef from butcheries in Kasama district

Following the Food and Agriculture Organisation (FAO) microbiological standards of fresh meat (Heinz and Hautzinger, 2007), it was reported that as regards the aerobic plate count 40.7% of butcheries sold meat in good bacterial condition (<4 Log10 cfu/g) while 40.7% and 18.5% were critical (4-5 Log10 cfu/g) and non-acceptable conditions (>5 Log10 cfu/g), respectively (Table 1). With the faecal coliform evaluation, 74.1% of the butcheries sold meat in good bacterial conditions (<2 Log10 cfu/g), while 14.8% and 11.1% were critical (2-3 Log10 cfu/g) and non-acceptable conditions (>3 Log10 cfu/g), respectively.

**Table 1:** Summary of Microbiological standards of fresh meat

|  | Number of Butcheries [Percent] |
| --- | --- |
| **Aerobic Plate Count Log10** |  |
| Good condition (Log10 <4 cfu/g) | 12 [40.7] |
| Critical condition (Log10 4-5 cfu/g) | 11 [40.7] |
| Not acceptable (Log10 >5 cfu/g) | 4 [18.5] |
| **Faecal Coliform Count Log10** |  |
| Good condition (Log10 <2 cfu/g) | 20 [74.1] |
| Critical condition (Log10 2-3 cfu/g) | 4 [14.8] |
| Not acceptable (Log10 >3 cfu/g) | 3 [11.1] |
| **Isolation pathogens** |  |
| *Salmonella* spp | 0 [0] |
| *Staphylococcus aureus* | 10 [37] |

From the meat samples analysed, Salmonella spp was not isolated from any of them while Staphylococcus aureus was isolated from 37% of the butcheries (Table 1).

### 3.2. Hygiene practices and management of food handlers in the butcheries

The butcheries in Kasama district purchased their beef carcasses from slaughter facilities within and outside the district. Results of the study revealed that 55.6% of the butcheries in Kasama district sourced beef carcasses from slaughter facilities outside the district while 33.3% purchased from the local slaughter slabs and 11.1% from both sources (Table 2).

**Table 2:** Summary of risk factors associated with meat quality in butcheries

| RISK FACTOR | RESPONSES |  |  |
| --- | --- | --- | --- |
| **Source of Beef Carcasses** | Within District [No. (%)] | Outside District [No. (%)] | Both |
|  | 9 (33.3) | 15 (55.6) | 3 (11.1) |
| **Cold storage facilities in butcherries** | Domestic deep freezer [No. (%)] | Walk in cold room [No. (%)] | Total [No. (%)] |
|  | 25 (92.6) | 2 (7.4) | 27 (100) |
| **Duration of beef storage in cold facilities (day)** | Mean | Min | Max |
|  | 11 | 1 | 30 |
| **Frequency of cleaning contact surfaces** | Mean | Min | Max |
|  | 7 | 1 | 15 |
| **Type of Cleaning Agent used** | Detergent [No. (%)] | Disinfectant [No. (%)] |  |
|  | 23 (85) | 4 (15) |  |

The two types of cold storage facilities recorded in the study were domestic deep freezer and walk in cold room which represented 92.6% and 7.4%, respectively (Table 2).

On average all butcheries stored beef in cold storage facilities for 11 days before it was purchased completely by consumers. The storage period ranged from 1 to 30 days (Table 2).

The average frequency of cleaning the meat contact surfaces such as tables, cutting surfaces and countertops was 7 times per day. The minimum frequency of cleaning was 1 and the maximum was 15(Table 2). The contact surfaces of the butcheries were cleaned using either detergents or disinfectants, the former being the most common (85%) and the latter being the least (15%), respectively.

## 4. DISCUSSION

The study revealed a varying level of bacterial contamination of the beef sold in butcheries of Kasama district. Aerobic plate count (APC) being a measure of microbial quality of the meat showed that 18.5% of the butcheries had beef that was in unacceptable conditions (>5 Log10 cfu/g), while 40.7% was in critical condition (4-5 Log10 cfu/g) and the remaining 40.7% was in good condition (<4 Log10 cfu/g). Presence of microbes in high numbers (APC >7 log CFU/cm2) fast tracks the spoilage of the meat. The high APC values were similar to the findings reported by Salihu et al (2013) in Nigeria and thereby predisposing the beef to easily spoilage (Salihu et al., 2013).

The study findings showed that 11.1 % of the butcheries had meat with faecal coliform counts of unacceptable levels (>3 Log10 cfu/g).These findings are in agreement with those of Datta et al., 2012 and Haque et al., 2008 who found high total coliform counts (>3 Log10 cfu/g) on sampled raw beef and goat meat respectively in Bangladesh (Datta et al., 2012; Haque et al., 2008). Therefore the results of the study suggest that this 11.1% of the beef that is sold in Kasama district is of poor bacterial quality and has a high likelihood of predisposing the consumers to foodborne infections. Concern should then be raised on the levels of hygienic practices during evisceration in the slaughtering process of beef carcasses, as the presence of faecal coliforms on the beef suggests that the puncture of gastrointestinal tract.

Staphylococcus aureus was isolated from beef in 37% of the sampled butcheries. Staphylococci are normal flora of the skin and upper respiratory tract in man and animals hence they can easily contaminate all foods types. If environmental conditions (e.g., time, pH, and temperature) during the production or storage of the food product are suitable for the growth of S. aureus, this microorganism produces enterotoxins which results in sub-acute gastroenteritis (Normanno et al., 2005). This finding is therefore a possible indicator of unhygienic handling of meat by the butchery personnel. It must be noted nonetheless, that low levels of staphylococci are expected to exist in all food products of animal origin or those that are directly handled by humans, unless heat processing is applied to effectively destroy them (Gracey and Collins, 1994). Staphylococcus aureus counts of 10^5^-10^6^ cells per gram are required to produce a pathogenic dose of enterotoxin that will lead to foodborne illness and the count of the bacteria was not determined in the study (Schelin et al., 2011).

A study of the beef value chain in Zambia revealed that larger meat processors transport carcasses from abattoirs in refrigerated trucks while small local butchers, however, transport meat from the abattoirs directly to the butchery in vans that lack refrigeration and sanitation (Lubungu et al., 2015). Therefore, the findings of the study indicating that 55.6% of butchers in Kasama purchased beef carcasses from abattoirs outside the district therefore reveals the likelihood of further contamination during transportation.

Proper refrigeration assures an inherent quality, while high temperature will exacerbate the quality deterioration related to spoilage by common psychotropic bacteria. From the study it was reported that the butcheries stored beef carcasses for an average of 11 days before they were completely sold off, which is longer than the recommended 3 to 5 days (Nychas et al., 2008). It was observed in the study that approximately 93% of the butcheries used domestic deep freezers for the storage of beef carcasses while only 7% had walk-in cold rooms. Light temperature fluctuation (+/- 5°) is normal in the freezer and refrigerator but this difference can be even greater depending on how frequently the door(s) is open and closed. A freezer that is constantly open cannot properly cool food. Temperature fluctuations during cold storage have been reported to promote the growth of psychrophiles and mesophiles (Dave and Ghaly, 2011). Even small increases of a degree or two can result in an enormous increase in bacterial growth (Gill and Gill, 2010). High meat surface temperatures not only encourage psychrotrophic bacteria to grow exponentially, accelerating the rate of discoloration and spoilage, but also provide ideal conditions for the growth of foodborne pathogens such as *Salmonella* (Gill and Gill, 2010). Therefore the use of domestic freezers for storage of beef may have an effect on the quality of the meat sold and its shelf life.

It was further observed that few butcheries (15%) used disinfectants to clean the contact surfaces compared to those who used various types of detergents (85%). Periodic cleaning and sanitation, which includes disinfection of butchery premises, counters and equipment is an integral part of good hygienic practices. These inadequate cleaning practices expose meat to contamination by pathogenic microorganisms, leading to adverse public health concerns.

## 5. CONCLUSION

The detection of *Staphylococcus aureus* and a seemingly high enumeration of both total and faecal coliforms in beef in butcheries and low hygiene standards raises concerns on the food safety management systems. The public health fraternity both from the local municipality and Ministry of health with the mandate to uphold food safety standards ought to engage all member of the beef value chain to ensure that the product being sold to consumers is wholesome and will not serve as vehicle for food poisoning.

## Data Availability

The authors confirm that the data supporting the findings of this study are available within the article [and/or] its supplementary material.

## 6. ACKNOWLEDGEMENT

We would like to recognize that this was a study undertaken by Ms. Sara Hanyinza as partial fulfilment for Bachelor of Science Environmental Health of Lusaka Apex Medical University (LAMU). We wish to express our gratitude to the staff at Kasama District Health Office, for technical support rendered during sample collection. We could like to thank the academic staff of LAMU, faculty of Health Sciences for the support and contribution towards the research. Finally, we are indebted to the butchery owners and their staff who allowed us to conduct the research in their premises, to them we say thank you.

